# Serum Calcium Level is Associated with Mortality Risk in Patients with Sepsis-induced Coagulopathy

**DOI:** 10.1101/2025.07.03.25330792

**Authors:** Qin Li, Yisong Cheng

## Abstract

**Background:** Electrolyte disorders are prevalent in patients with sepsis-induced coagulopathy (SIC). The aim of this study was to explore the association between serum calcium levels and mortality in patients with SIC.

**Methods:** We screened the Medical Information Market for Intensive Care III database for the data of patients with SIC. Logistic regression analysis was performed to explore the risk factors for mortality in patients with SIC, and restricted cubic spline (RCS) was applied to fit the correlation between the serum calcium levels and SIC mortality.

**Results:** The in-hospital mortality of patients with SIC was 32.2%. The mortality rate of patients in the first and third tertiles of serum calcium levels was higher than that of patients in the second tertile of serum calcium levels. Results of the univariate regression analysis revealed that serum calcium levels were not associated with mortality. The unadjusted RCS suggested a U-shaped relationship between serum calcium level and mortality. After adjusting for confounding factors, multivariate regression analysis revealed that the relationship between serum calcium level and mortality remained U-shaped.

**Conclusion:** The incidence of mortality was high among patients with SIC who had abnormally low or high serum calcium levels. Physicians should pay attention to the clinical management of patients with SIC, especially hypercalcemia.

## 1. Introduction

Sepsis-induced coagulopathy (SIC) is caused by vascular endothelial cell injury and coagulation dysfunction caused by sepsis. The International Society on Thrombosis and Haemostasis (ISTH) proposed its diagnostic criteria in 2017 [1]. Globally, 24.0–60.0% of patients with sepsis have SIC, compared to 67.9% in China. Improper management can lead to the development of disseminated intravascular coagulation (DIC), which increases patient mortality by twofold [2–6]. Although many patients with SIC survive aggressive treatment, they draw attention due to the significant burden of prolonged hospital and intensive care unit (ICU) stays and high healthcare costs for the families.

SIC is a coagulation-related disorder caused by the effect of sepsis activation on the vascular endothelium and the effects of inflammation, immunity, and other factors on the coagulation system [1]. In addition to routine coagulation testing, biomarkers of the coagulation pathway can be used for early prediction of SIC [7]. Calcium ions are electrolytes and important coagulation factors in endogenous and exogenous coagulation pathways. Calcium ion level can be easily assessed with high accuracy; therefore, it has potential research value.

Calcium disorders are common among critically ill patients, and studies have explored the prognostic value of serum calcium levels in these patients. Hypocalcaemia has been shown to be associated with increased mortality in critically ill patients [8–11]. Li et al. reported that serum calcium levels are valuable in the early identification and severity of sepsis in elderly patients [12]. Another study showed that serum calcium levels in patients with sepsis were associated with a U-shaped 28-day mortality rate, and hypercalcaemia and hypocalcaemia were associated with an increased risk of 28-day mortality in patients with sepsis [13]. However, to our knowledge, no clinical study has evaluated the correlation between serum calcium levels and SIC. Therefore, we aimed to retrospectively analyse the relationship between serum calcium levels and SIC in patients with sepsis admitted in the ICU.

## 2. Materials and Methods

### 2.1 Patients

Research data were obtained from the Medical Information Market for Intensive Care (MIMIC)-III database that includes unidentified health-related data of ICU hospitalisations at the Beth Israel Deacons Medical Center (BIDMC) between June 2001 and October 2012; it was constructed by researchers at the Massachusetts Institute of Technology (MIT) Computational Physiology Laboratory and a collaborative research team. The project was approved by the MIT and BIDMC Institutional Review Boards and granted an exemption for informed consent. All adults who met the diagnosis of SIC were included.The exclusion criteria were as follows: (1) age <18 years and (2) lack of vital signs and laboratory examination records (Figure 1). After application of the inclusion and exclusion criteria, 6889 patients were included in the analysis. The study complied with the ethical standards of the Declaration of Helsinki.

**Figure 1.**
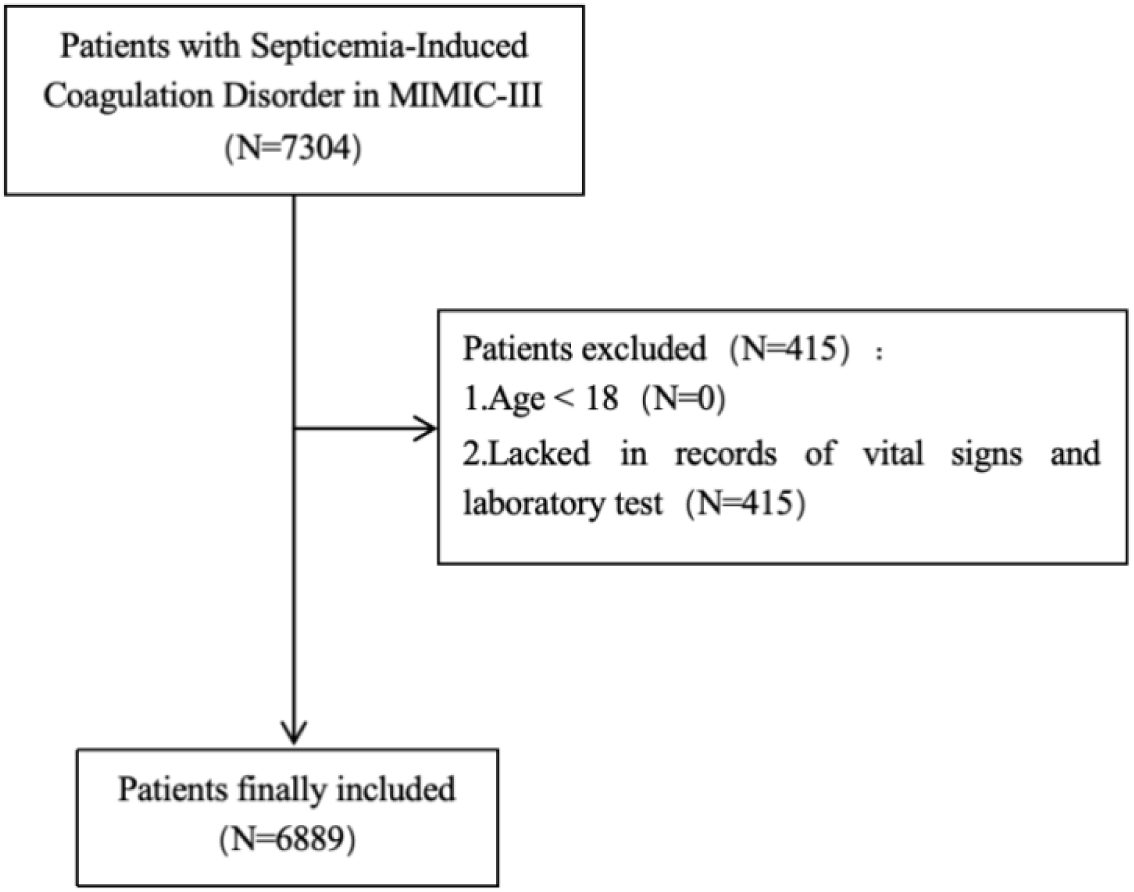
Flowchart of patients inclusion. Medical Information Mart for Intensive Care—III (MIMIC-III).

### 2.2 Data Collection

The basic data of all patients was extracted from the MIMIC-III database. Baseline characteristics included sex, age, and six comorbidities: diabetes, hypertension, myocardial infarction, congestive heart failure, chronic liver disease, and chronic kidney disease. Vital signs included systolic blood pressure, diastolic blood pressure, heart rate, and oxygen saturation. Data from the following laboratory tests after SIC diagnosis were extracted: white blood cell (WBC); platelet; red blood cell (RBC) counts; haematocrit (HCT); and haemoglobin, blood urea nitrogen, serum creatinine, blood glucose, blood calcium, blood potassium, and blood sodium levels. Data on in-hospital mortality were also recorded. All variables were extracted from the MIMIC-III database using Navicat Premium 12 Structure Query Language.

### 2.3 Statistical Analysis

The Kolmogorov–Smirnov test was used to determine the normality of the recorded variables. Normally and non-normally distributed variables were expressed as means ± standard deviations and medians (interquartile ranges), respectively. Categorical variables were presented as numbers (percentages). The t-test and Mann–Whitney U test were used to compare the differences in the normally and non-normally distributed variables between the two groups.

Chi-square test or Fisher’s exact test was used to compare the differences in categorical variables between the two groups. A restricted cubic spline (RCS) was applied to determine the association between serum calcium levels and mortality risk. Univariate and multivariate logistic regression analyses were used to analyse the risk factors for mortality and to identify the relationship between serum calcium level and mortality in patients with SIC. Multiple regression analysis was used for the subgroup analysis. Statistical significance was defined as a two-sided p-value <0.05. IBM SPSS Statistics for Windows, version 26.0 (IBM Corp., Armonk, NY, USA), and R software (version 3.6.1; R Foundation for Statistical Computing, Vienna, Austria) were used for all statistical analyses and figure illustrations.

## 3. Results

### 3.1 Baseline Characteristics of Patients with SIC

Among the 6889 enrolled patients with SIC, 2225 died, resulting in an in-hospital mortality rate of 32.3% (Table 1). Non-survivors were older (*p* < 0.001); had a history of myocardial infarction (*p* < 0.001); and had a higher incidence of congestive heart failure (*p* = 0.041), chronic liver disease (*p* < 0.001), and chronic renal disease (*p* < 0.001) than the survivors. Further, blood urea nitrogen (*p* < 0.001), serum creatinine (*p* < 0.001), glucose *(p* < 0.001), and potassium (*p* < 0.001) levels were higher among the non-survivors, while calcium (*p* < 0.001), sodium (*p* = 0.013) and platelet (*p* < 0.001) were lower in them compared to the survivors. Serum calcium levels were significantly different between survivors and non-survivors (*p* < 0.001). The mean serum calcium level was 7.69 mg/dL (SD: 0.93). The serum calcium level showed normal distribution, with the highest distribution between 7.4 and 8.2 mg/dL (Figure 2). Patients were divided into three groups according to tertiles of serum calcium levels (≤7.4 mg/dL, 7.5–8.0 mg/dL, ≥8.1 mg/dL). The mortality rates in the first, second, and third tertiles were 34.4%, 29.0%, and 32.7%, respectively (Figure 3). Additionally, the length of ICU stay was longer (*p* = 0.014) and the length of hospital stay was shorter among the non-survivors compared to the survivors *(p* < 0.001).

**Figure 2.**
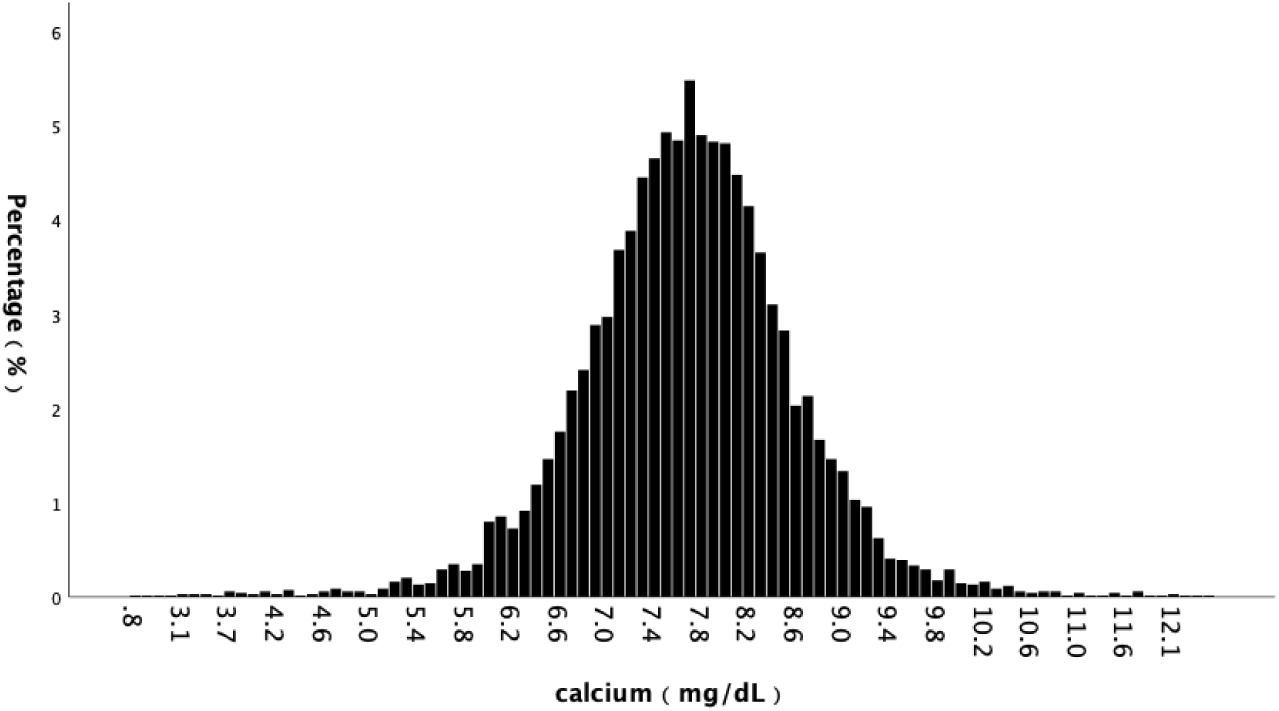
Distribution of minimal serum calcium level in included SIC patients.

**Figure 3.**
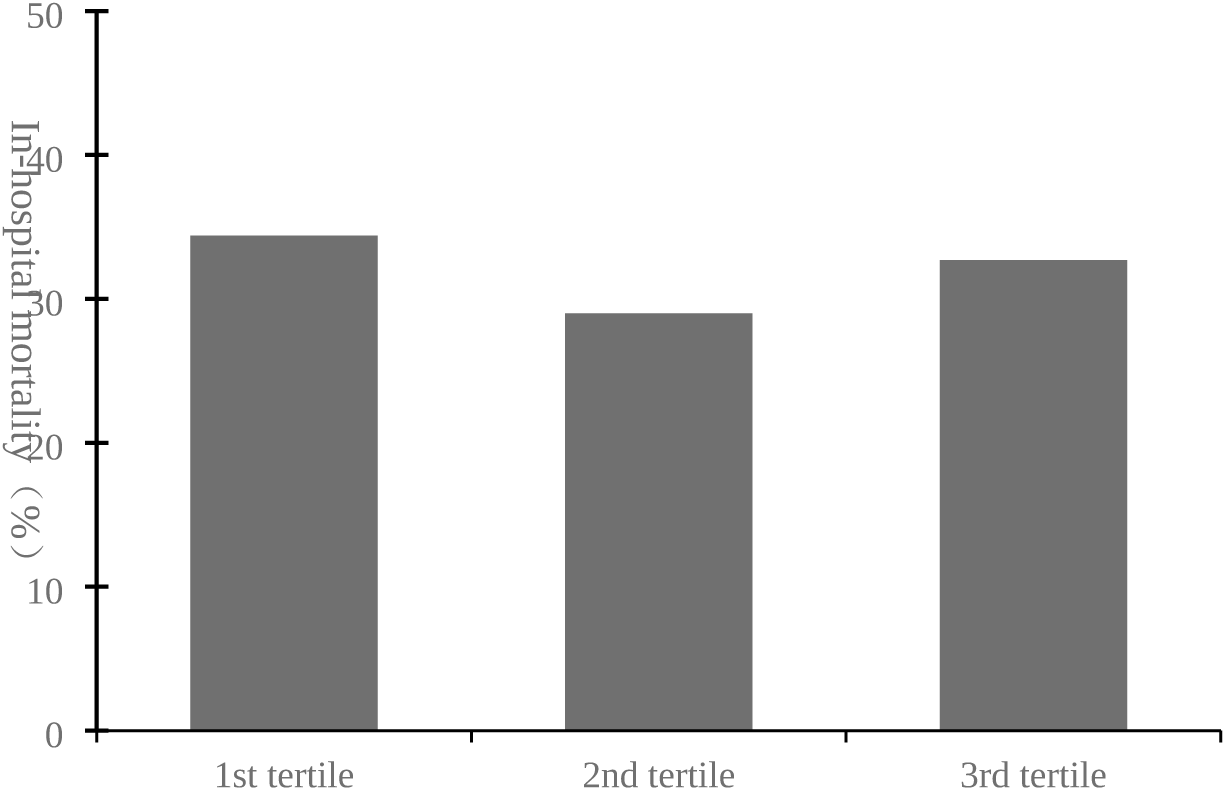
Mortality of three groups devided by serum calcium level.

**Table 1.**
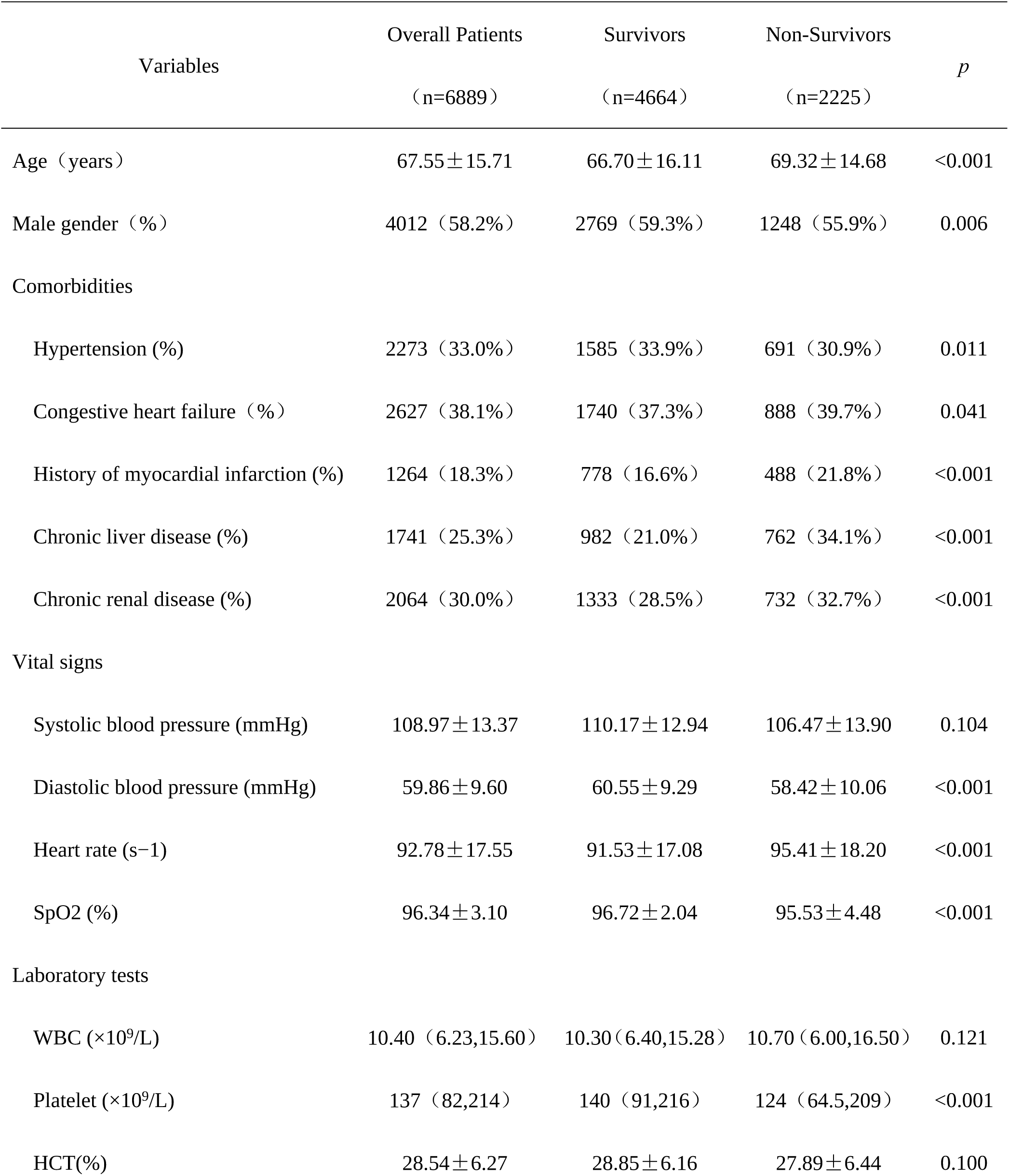
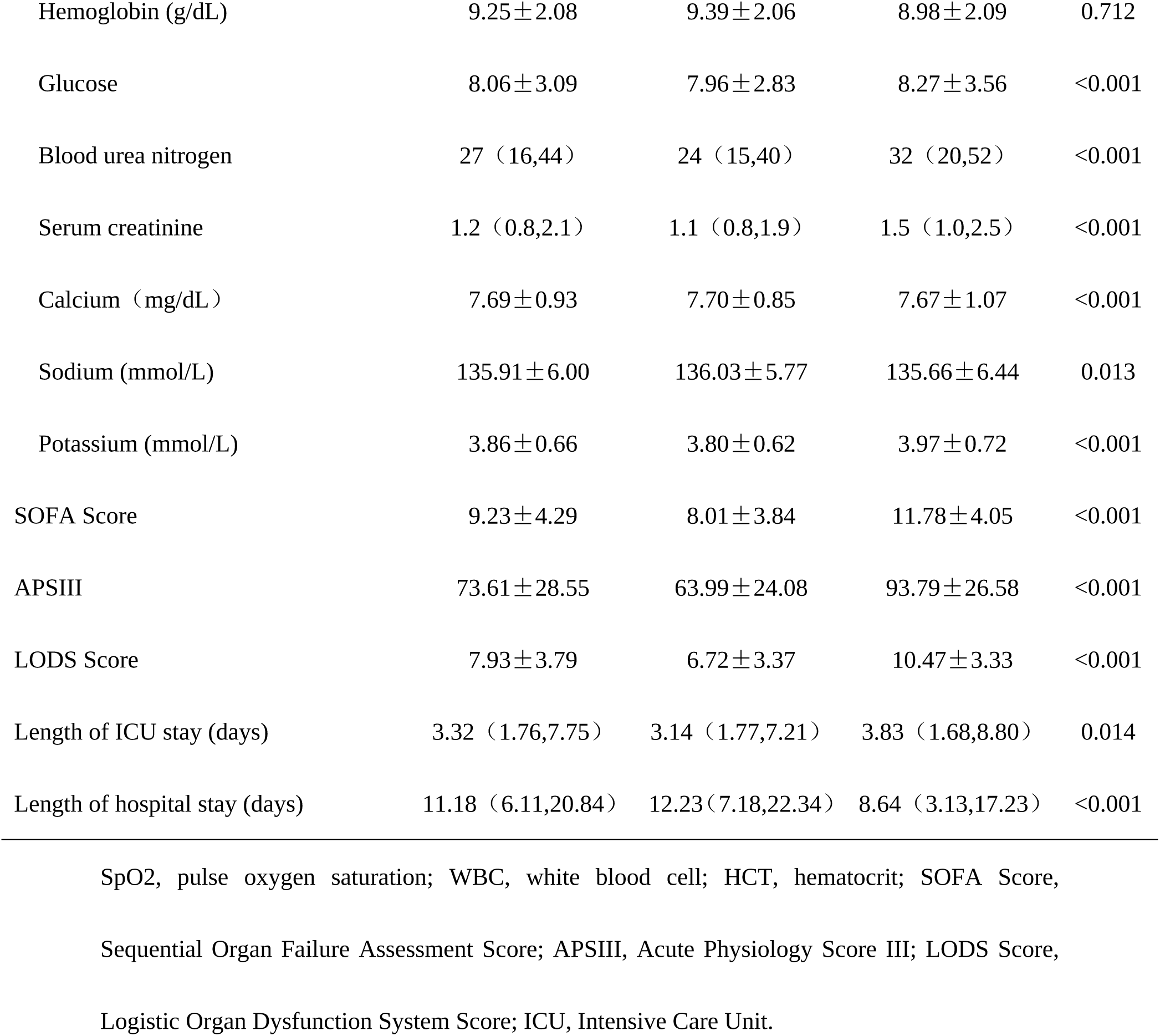
Baseline characteristics of included SIC patients.

### 3.2 Unadjusted Association between Serum Calcium Level and Risk of Mortality

Univariate logistic regression indicated that sex (*p* = 0.006), age (*p* < 0.001), hypertension (*p* = 0.012), history of myocardial infarction (*p* < 0.001), congestive heart failure (*p* = 0.041), chronic renal disease (*p* < 0.001), chronic renal disease (*p* < 0.001), heart rate (*p* < 0.001), systolic blood pressure (*p* < 0.001), diastolic blood pressure (*p* < 0.001), SpO_2_ (*p* < 0.001), WBC (*p =* 0.003), haemoglobin (*p* < 0.001), platelet (*p* < 0.001), HCT (*p* < 0.001), blood urea nitrogen *(p* < 0.001), serum creatinine (*p* < 0.001), glucose (*p* < 0.001), sodium (*p* = 0.017), and potassium (*p* < 0.001) were potential risk factors of in-hospital mortality among patients with SIC (Table 2). A significant association was observed between serum calcium level and mortality risk (*p* < 0.001). Unadjusted RCS curve showed a U-shaped nonlinear relationship between serum calcium levels and mortality, with the lowest mortality in patients with serum calcium levels between 8.8 and 10.8 mg/dL, consistent with the second tertile (Figure 4A).

**Figure 4.**
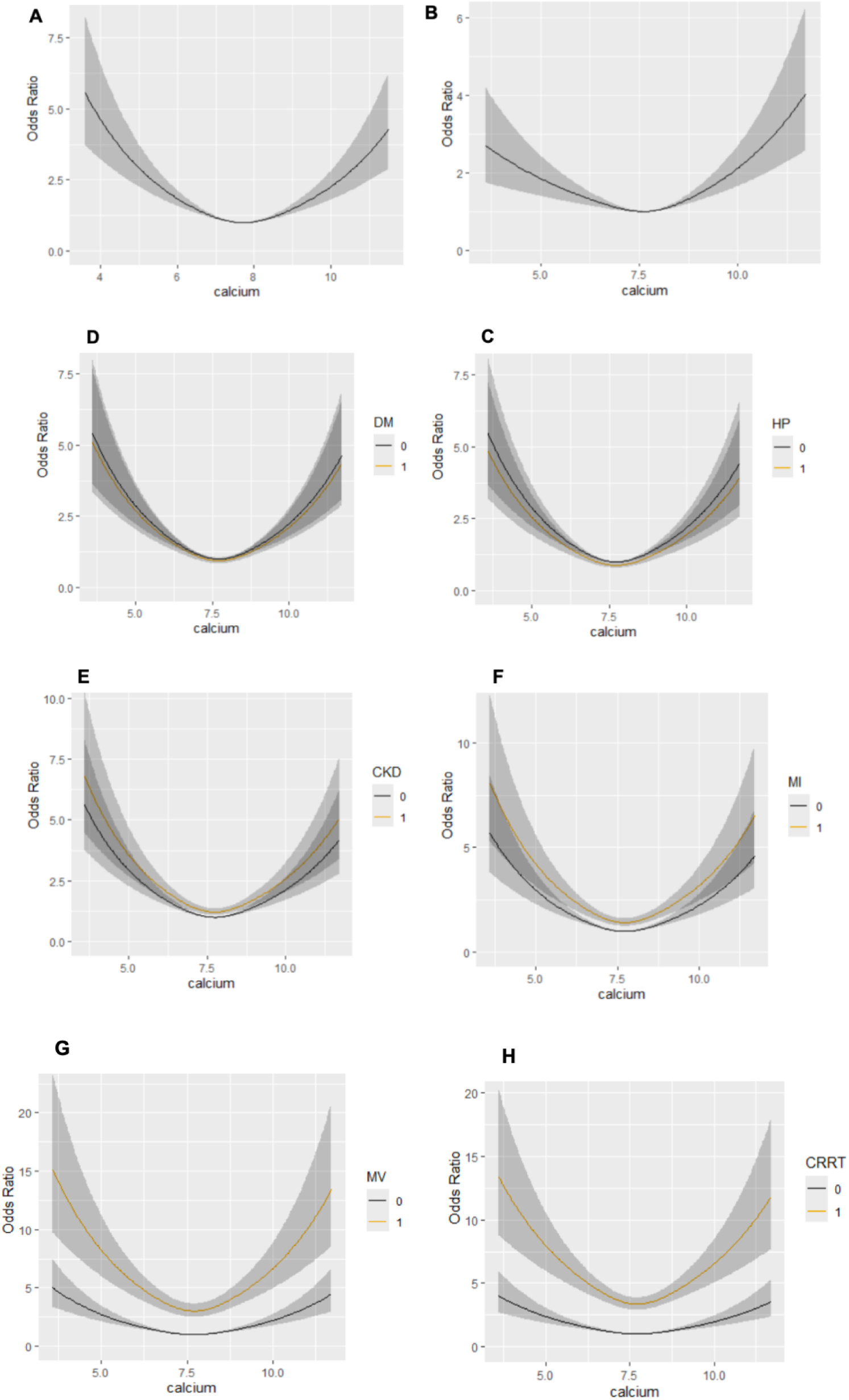
**(A)** Unadjusted association between serum calcium level and risk of mortality. **(B-H)** Adjusted association between serum calcium level and risk of mortality. Adjusted for gender, hypertension, diabetes, history of myocardial chronic kidney disease, history of myocardial infarction, ventilation, CRRT, heart rate, SBP, DBP, SPO_2_, WBC, platelets, hematocrit, hemoglobin, bun, creatinine, sodium, potassium, length of ICU stay, length of hospital stay.

**Table 2.** Univariate logistic regression analysis of risk factors for coagulopathy in SIC patients.

| Variables | OR | 95% CI | *p* |
| --- | --- | --- | --- |
| Age | 1.011 | 1.008-1.014 | <0.001 |
| Gender | 0.866 | 0.782-0.959 | 0.006 |
| Diabetes | 0.937 | 0.842-1.042 | 0.230 |
| Hypertension | 0.870 | 0.780-0.969 | 0.012 |
| Congestive heart failure | 1.114 | 1.004-1.236 | 0.041 |
| History of myocardial infarction | 1.396 | 1.230-1.584 | <0.001 |
| Chronic liver disease | 1.941 | 1.735-2.172 | <0.001 |
| Chronic renal disease | 1.223 | 1.097-1.363 | <0.001 |
| Systolic blood pressure | 0.978 | 0.974-0.982 | <0.001 |
| Diastolic blood pressure | 0.976 | 0.971-0.982 | <0.001 |
| Heart rate | 1.013 | 1.010-1.016 | <0.001 |
| SpO<sub>2</sub> | 0.873 | 0.856-0.891 | <0.001 |
| WBC | 1.008 | 1.003-1.013 | 0.003 |
| Platelet | 0.999 | 0.998-0.999 | <0.001 |
| HCT | 0.975 | 0.967-0.983 | <0.001 |
| Hemoglobin | 0.908 | 0.886-0.931 | <0.001 |
| Glucose | 1.031 | 1.015-1.048 | <0.001 |
| Blood urea nitrogen | 1.013 | 1.011-1.015 | <0.001 |
| Serum creatinine | 1.110 | 1.076-1.145 | <0.001 |
| Calcium |  |  | <0.001 |
| Calcium (1 vs. 2) | 1.192 | 1.047-1.357 | 0.008 |
| Calcium (3 vs. 2) | 1.283 | 1.132-1.454 | <0.001 |
| Sodium | 0.990 | 0.981-0.998 | 0.017 |
| Potassium | 1.467 | 1.359-1.585 | <0.001 |
| Length of ICU stay | 1.008 | 1.002-1.014 | 0.006 |
| Length of hospital stay | 0.986 | 0.982-0.990 | <0.001 |

OR, odds ratio; CI, confidence interval; SpO<sub>2</sub>, pulse oxygen saturation; WBC, white blood cell; HCT, hematocrit.

### 3.3 Adjusted Association between Serum Calcium Level and Risk of Mortality

After adjusting for the confounding factors, in a segmented regression model based on serum calcium level and the risk of in-hospital mortality, the RCS curve suggested that the cut-off point of serum calcium was 7.5 mg/dL (Figure 4B). In addition, a U-shaped relationship between serum calcium and in-hospital mortality was observed for patients with or without hypertension, diabetes, chronic kidney disease, and myocardial infarction, and patients with or without mechanical ventilation and continuous renal replacement therapy (Figure 4C-H). Multiple logistic regression analysis confirmed this relationship (Table 3).

**Table 3.** Multivariate logistic regression analysis of risk factors for coagulopathy in SIC patients.

|  | OR | 95% CI | *p* |
| --- | --- | --- | --- |
| Model 1 |  |  |  |
| Calcium (1 vs. 2) | 1.132 | 0.991-1.293 | 0.068 |
| Calcium (3 vs. 2) | 1.364 | 1.199-1.551 | <0.001 |
| Model 2 |  |  |  |
| Calcium (1 vs. 2) | 1.206 | 1.051-1.384 | 0.008 |
| Calcium (3 vs. 2) | 1.209 | 1.058-1.382 | 0.005 |
| Model 3 |  |  |  |
| Calcium (1 vs. 2) | 1.159 | 1.007-1.334 | 0.039 |
| Calcium (3 vs. 2) | 1.213 | 1.057-1.392 | 0.006 |

OR, odds ratio; CI, confidence interval. Model 1 adjusted for age, diabetes, hypertension, myocardial infarction, congestive heart failure, chronic liver disease, chronic renal disease; Model 2 adjusted for age, diabetes, hypertension, myocardial infarction, congestive heart failure, chronic liver disease, chronic renal disease, sbp, dbp, heart rate, spo<sub>2</sub>; Model 3 adjusted for age, diabetes, hypertension, myocardial infarction, congestive heart failure, chronic liver disease, chronic renal disease, sbp, dbp, heart rate, spo<sub>2</sub>, glucose, hematocrit, hemoglobin, platelets, wbc, bun, creatinine, sodium, potassium.

## 4. Discussion

In this retrospective cohort study, the data of patients with SIC showed a U-shaped association between serum calcium levels and in-hospital mortality. Serum calcium levels of <7.5 mg/dL and >7.5 mg/dL were negatively and positively correlated with the in-hospital mortality risk, respectively. Serum calcium levels close to 7.5 mg/dL were associated with the lowest risk of in-hospital mortality in patients with SIC.

Wang et al. described a U-shaped relationship between the initial total calcium level and hospital mortality in critically ill patients [14]. Both hypercalcaemia and hypocalcaemia were associated with increased hospital mortality when adjusted for clinical characteristics, including age and sex. In many previous studies, ionized calcium was associated with mortality among patients with critical illness [15–16]; however, the relationship between serum calcium levels and mortality has not been studied. To the best of our knowledge, this study is the first to establish an association between serum calcium levels and mortality in patients with SIC.

A variety of risk factors, such as unfractionated heparin anticoagulant therapy [17] and serum sodium [18], serum zinc [19], plasma infusion [20], thrombocytopenia [21], and C1q [22] levels have been associated with poor prognosis in patients with SIC. Consistent with previous studies, the results of the univariate analysis in this study showed that age; history of heart disease, liver disease, and kidney disease; decreased SBP, DBP, and SPO_2_; increased heart rate; low platelet count; and increased urea nitrogen and creatinine levels were positively correlated with in-hospital mortality in SIC. Therefore, early intervention and management strategies are needed for patients with SIC and these risk factors to improve their prognosis.

SIC ranges from moderate immune thrombosis to excessive thrombus inflammation. An immune thrombus is formed in microvessels in the early stages of infection and has an immune function. Excessive formation of immune thrombi can lead to disorders and spread of thrombus inflammation, leading to SIC [23]. Thrombosis is an innate immune response involving white blood cells and platelets. During sepsis, activated neutrophils release extracellular traps to limit infection. These traps are composed of DNA, histones, and other neutrophil granule proteins, all of which are highly prothrombotic [24]. After neutrophil death, DNA and histones are released to form neutrophil extracellular traps (NETs). NETs, platelets, and fibrinogen form immune thrombi that intercept, separate, capture, and kill pathogens. Viable neutrophils can also excrete histones and mitochondrial DNA in a non-cytolytic manner and participate in the formation of NETs [25].

The antithrombotic effect of the vascular endothelium is also important in sepsis; however, it can prevent clot formation, maintain vascular integrity, and regulate vascular tension under physiological conditions. Vascular endothelial cells release nitric oxide and prostacyclin, which contribute to physiological anti-thrombosis. Under septic conditions, it promotes thrombosis by expressing tissue factors and releasing the von Willebrand factor. When the systemic immune response continues, the extensive formation of immune microthrombi can lead to uncontrolled thrombotic inflammation, aggravate vascular endothelial cell dysfunction, cause capillary leakage and consumptive coagulopathy, and even cause DIC and multiple organ failure [26].

The specific physiological mechanisms related to serum calcium levels and the prognosis of SIC patients are still unclear. Moe SM has reported that parathyroid hormones, albumin, vitamin D (1,25-dihydroxyvitamin D or 25-hydroxyvitamin D), and other factors can regulate calcium levels [27]. In sepsis, pro-inflammatory cytokines IL-6 and IL-1β can upregulate the expression of parathyroid and renal calcium-sensitive receptors, ultimately reducing blood calcium levels [28]. In contrast, dysregulation of serum calcium levels may activate calcium-sensitive receptors on the surface of T cells, promote ROS and cytokine release, impairing endothelial cell and barrier function, ultimately leading to fluid leakage, tissue inflammation, and adverse prognosis in sepsis [29]. Calcium is a key cofactor of the coagulation cascade. Improper blood calcium levels can impair platelet function, affect the coagulation cascade, and lead to bleeding events and adverse prognosis in sepsis patients [30–31]. This may explain the correlation between hypocalcaemia and a poor prognosis in patients with SIC.

Our study had some limitations. First, this was a single-centre, retrospective, cohort study; therefore, selection bias was inevitable. However, we adjusted for as many potential confounders as possible in the data analysis to minimise potential bias. Secondly, it is difficult to determine the causal relationship and mechanism between calcium disorders and adverse outcomes in SIC patients. Further experimental and prospective studies are still needed to explore the mechanism and key role of serum calcium levels in SIC prognosis. Third, we only analysed the relationship between minimal calcium levels and SIC prognosis during hospitalization, while the relationship between dynamic trends in serum calcium levels and SIC prognosis is not yet clear. Finally, our endpoint indicator was in-hospital mortality, and the association between serum calcium levels and the long-term prognosis of patients with SIC was not explored.

## 5. Conclusions

Both hypocalcaemia and hypercalcaemia were associated with high mortality rates in patients with SIC. Our results indicated that hypercalcemia levels are independently associated with death in SIC patients. Clinicians should pay attention to the management of SIC patients, especially those with high serum calcium.

## Author Contributions

Q. L. and Ys. C. conceived the project and drafted the manuscript. Q.L. performed data extraction, analysed the data, and drafted the manuscript. Ys. C. has made strict revisions to the knowledge content of the manuscript. All authors have read and agreed to the publication.

## Institutional Review Board Statement

The data for this study are from the MIMIC database established by the Beth Israel Deaconess Medical Center (BIDMC). The database was approved by the Institutional Review Board of MIT and BIDMC. To protect privacy, all patients in this database are anonymous.

## Informed Consent Statement

Patient informed consent was exempted due to the retrospective design of this study.

## Data Availability Statement

The data set is available from the corresponding author upon request.

